# The impact of assortative mating, participation bias, and socioeconomic status on the polygenic risk of behavioral and psychiatric traits

**DOI:** 10.1101/2022.11.29.22282912

**Authors:** Brenda Cabrera-Mendoza, Frank R Wendt, Gita A Pathak, Loic Yengo, Renato Polimanti

## Abstract

To investigate assortative mating (AM), participation bias, and socioeconomic status (SES) with respect to the genetics of behavioral and psychiatric traits, we analyzed gametic phase disequilibrium (GPD), within-spouses and within-siblings polygenic risk score (PRS) correlation, performing a SES conditional analysis. We observed genetic signatures of AM across multiple methods for traits related to substance use with SES conditioning increasing the within-spouses PRS correlation for *Frequency of drinking alcohol* (2.5% to 6%), *Maximum habitual alcohol intake* (1.33% to 4.43%), and *Ever taken cannabis* (1.5% to 5.3%). Comparing UK Biobank mental health questionnaire responders vs. non-responders, major depressive disorder PRS showed significant GPD in both groups when based on the Million Veteran Program (3.2% vs. 3%), but only in responders when based on the Psychiatric Genomics Consortium (3.8% vs. 0.2%). These results highlight the impact of AM, participation bias, and SES on the polygenic risk of behavioral and psychiatric traits.

## Introduction

Psychiatric disorders and traits are highly polygenic, i.e. they are influenced by several thousands of genetic variants, each having a small effect on disease risk^1^. Large-scale genome-wide studies have demonstrated that our ability to investigate their polygenic architecture could be influenced by several factors such as assortative mating (AM; i.e., mate choice driven by phenotypic similarity) and participation bias (i.e., individuals with a certain phenotype are more likely to enter a study)^2-4^. For example, AM increases the genetic variance in a population because it induces a systematic positive correlation between trait-increasing alleles across the genome^5^. Therefore, AM can result in inflated genetic effects as compared to those estimated in randomly mating population or using a family-based design. On the other hand, participation bias might exacerbate differences between sub-groups in a study, thereby reducing the generalizability of genetic effects estimated from the whole sample^6^.

As exemplified hereafter, both AM and participation bias have been linked to behavioral traits and psychiatric disorders. A previous study investigating more than 700,000 individuals reported evidence of mate resemblance within and across eleven psychiatric disorders, including attention-deficit/hyperactivity disorder (ADHD), autism spectrum disorder (ASD), schizophrenia, bipolar disorder, major depressive disorder (MDD), generalized anxiety disorder (GAD), agoraphobia, social phobia, obsessive-compulsive disorder (OCD), anorexia, and substance abuse^7^. A subsequent analysis provided an estimate of the genetic consequences of AM for psychiatric traits, suggesting a modest impact on their heritability but this may be considerable for the population prevalence of rare disorders with a high heritability^8^. Recently, an analysis of cross-phenotype AM highlighted how cross-phenotype mate correlations may bias estimates of genetic correlation between pairs of psychiatric disorders^9^.

With respect to participation bias^10-15^, certain psychiatric traits (e.g., MDD, anxiety, and alcohol consumption) can be associated with the likelihood of becoming or remaining as study participants^16,17^. Genome-wide investigations showed that the non-participation in health surveys (including mental health assessments) is genetically correlated with several behavioral traits, such as educational attainment and neuroticism, and neuropsychiatric disorders, such as schizophrenia and Alzheimer’s disease^10,13^. Additionally, differential participation bias between sexes (i.e., participation bias where the determinants of study participation affect women and men to differing extents) is genetically correlated with behavioral and psychiatric traits, including educational attainment, risk-taking behaviors, cannabis use, loneliness, MDD, ASD, schizophrenia, and ADHD^12^.

Factors responsible for AM (e.g., opportunities or exposure to potential intimate partners, individual preferences, and third-party constraints) can be strongly affected by socioeconomic status (SES)^18^. Similarly, the proportions of non-participation are typically not uniform across sociodemographic groups, with those from deprived backgrounds often under-represented in health surveys^15^. To our knowledge, no previous study systematically investigated the potential impact of SES on the genetic signatures of AM and participation bias across multiple behavioral and psychiatric traits. Accordingly, we evaluated whether the SES association with the polygenic risk of psychiatric and behavioral traits contributes to the genetic signatures of AM and participation bias. Specifically, we leveraged findings from large-scale genome-wide association studies generated by the Psychiatric Genomics Consortium (PGC)^19^ and the Million Veteran Program (MVP)^20^ together with individual-level data from UK Biobank (UKB) participants that completed the UKB Mental Health Questionnaire (MHQ) and comparing to those that did not complete this assessment.

## Methods

### UK Biobank

The UKB is a general population-based cohort comprising approximately 502,000 participants. This sample was recruited between 2006 and 2010 in 22 assessment centers across the UK^21^. UKB received ethical approval from the NHS National Research Ethics Service Northwest (reference: 11/NW/0382). UKB obtained informed written consent from all participants. A self-reported detailed account of sociodemographic, lifestyle, mental, and physical health information was collected from all UKB participants^21^.

The collection and processing of UKB genetic data have been described previously^22^. Briefly, genome-wide genotype data were obtained from all UKB participants using the UKB Axiom array. UKB genotypic data were imputed using the Haplotype Reference Consortium reference panel. In this study, we analyzed a sample of 362,132 unrelated individuals of European descent (EUR) with available genotype data. Because of the limited sample size available, we were not able to analyze other ancestry groups. Ancestry and relatedness information of each UKB participant were obtained from the Pan-ancestry genetic analysis of the UKB (Pan-UKB)^23^. Briefly, genetic relatedness among UKB participants was estimated with PC-Relate, a principal component analysis (PCA)-based method^24^. While ancestry assignment was performed using a combined reference panel including both the 1000 Genomes Project^25^ and Human Genome Diversity Panel (HGDP)^26^. Then, a random forest classifier trained with the top six principal components (PCs) from the reference data was applied to the UKB PCs data. UKB participants were assigned to an ancestry group (African, Admixed American, Central/South Asian, East Asian, EUR, or Middle Eastern) based on a classifier probability >50%. A detailed description of the Pan-UKB methods is available at https://pan.ukbb.broadinstitute.org.

### UKB Mental Health Questionnaire

As part of the UKB assessment, behavioral and psychiatric outcomes, including mood disorders, anxiety, mental distress, self-harm, traumatic events, substance use, and post-traumatic stress disorder (PTSD), were evaluated with an online follow-up assessment including the UKB MHQ (Resource 22 on http://biobank.ctsu.ox.ac.uk)^27^. The MHQ was completed by 157,366 UKB participants (31% of total UKB participants) aged 45–82 years, 57% of them were female and had a higher SES (i.e., higher income and higher educational attainment) compared with UKB participants who did not complete this assesment^27,28^.

Also, diagnostic criteria were evaluated for MDD, hypomania or mania, GAD, alcohol use disorder (AUD), and PTSD^27^. Addiction to substances other than alcohol and/or behavior was defined based only on self-report. The MHQ is based on previously existing and validated measures, including the Composite International Diagnostic Interview Short Form (CIDI-SF) to assess lifetime mental disorders in general^29^, as well as instruments for specific mental disorders and trauma exposures, i.e., the PTSD Check List - Civilian Short version (PCL-6), the Childhood Trauma Screener – 5 item (CTS-5)^30,31^, and validated instruments developed specifically for the UKB such as an adult trauma screener^27^.

### Large-scale Genome-wide Association Studies of Behavioral and Psychiatric Traits

To investigate additional behavioral and psychiatric traits, we investigate AM genetic signatures using GWAS statistics that were generated from samples that did not include UKB. These included ADHD^32^, ASD^33^, anorexia nervosa^34^, anxiety disorder^35^, bipolar disorder^36^, bipolar disorder type I^36^, bipolar disorder type II^36^, MDD^37^, schizophrenia^38^, panic disorder^39^, PTSD^40^, Tourette syndrome^41^, and OCD^42^ obtained from the PGC (available at https://pgc.unc.edu/for-researchers/download-results/); and AUD^43^, Alcohol Use Disorder Identification Test-Consumption (AUDIT-C)^43^, maximum habitual alcohol intake^44^, MDD^45^, opioid use disorder (OUD)^46^, PTSD^47^, and anxiety disorder^48^ obtained from the MVP (available at https://www.ncbi.nlm.nih.gov/projects/gap/cgi-bin/study.cgi?study_id=phs001672.v8.p1). Briefly, PGC data was generally generated from meta-analyses of genome-wide genetic data derived from many cohorts with different characteristics and assessed with different instruments^49^. Conversely, MVP data were obtained from a single observational cohort study of US veterans^50^. The genetic correlation between PGC and MVP overlapping traits, i.e., MDD, PTSD, and anxiety was calculated with the Linkage Disequilibrium Score Regression method (LDSC)^51^. A description of these datasets is shown in Supplementary Table 1. Due to the limited sample size available for other ancestry groups in UKB, PGC, and MVP, our analyses were limited only to EUR.

### Gametic Phase Disequilibrium Analysis

We estimated AM genetic signatures across behavioral and psychiatric traits using a method proposed by Yengo et al.,^5^. Briefly, this method is based on the fact that AM signatures of a specific trait can be quantified as the directional correlation between trait-increasing alleles, also referred as gametic phase disequilibrium (GPD). This can be estimated as the correlation between trait-specific PRS based on variants located on odd and even chromosomes^5^.

Initially, we used GWAS statistics generated from the analysis of MHQ traits assessed in EUR MHQ responders to calculate PRS with respect to 243,476 EUR unrelated UKB participants who did not respond to the MHQ (MHQ non-responders). MHQ GWAS statistics were derived from the Pan-UKB analysis that used a generalized mixed model association testing framework available from the Scalable and Accurate Implementation of GEneralized (SAIGE) software^52^. A detailed description of these GWAS is available at https://pan.ukbb.broadinstitute.org. To reduce the multiple-testing burden, we tested only MHQ traits with SAIGE heritability estimates > 0.03 and single nucleotide polymorphisms-based heritability (SNP-h^2^) p-value < 0.05. For each MHQ trait, SNP-h^2^ was estimated using the LDSC approach^51^ as described in the Supplemental Methods.

Before PRS calculation, quality control was performed on GWAS summary statistics and UK individual genotypic data using PLINK 1.9^53^. SNPs were included if they meet the following criteria: i) Hardy– Weinberg equilibrium (HWE) p values >1 × 10^−6^, ii) missingness <0.05, iii) minor allele frequencies ≥ 0.01. PRS analysis included only LD-independent SNPs, selected with a clump r2<0.1 for SNPs < 1Mb apart using 1000 Genomes Project EUR populations as reference^25^.

Even- and odd-chromosomes PRS were calculated using the software package PRSice^54,55^. We included 20 PCs from SNPs in even and odd chromosomes when calculating the PRS for odd and even chromosomes, respectively to correct for population stratification. PCs were calculated from LD-independent SNPs in even and odd chromosomes separately using the fast PCA approach implemented in PLINK version 2.0^56,57^. LD pruning was performed in PLINK (r2<0.1 for SNPs < 1Mb apart) using HapMap3 (Utah residents with ancestry from northern and western EUR (CEU) as reference^58^.

As suggested by the GPD method developers^5^, a P-value threshold of 0.005 was applied to select SNPs included for PRS calculation. Thus, AM was estimated as the coefficient from a linear regression model of the even-chromosomes PRS (PRS_even_) onto odd-chromosomes PRS (PRS_odd_) and 20 PCs from the SNPs in odd chromosomes:

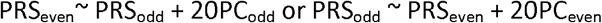

AM estimate of each MHQ trait was obtained from the regression onto the PRS with the larger variance. A false discovery rate (FDR q⍰<⍰0.05) was applied to correct the GPD results for the number of phenotypes evaluated in each analysis. Then, we calculated GPD estimates for psychiatric traits in unrelated EUR UKB participants (N=362,132), and to assess the effect of SES on participation bias in the UKB, we compared GPD estimates for psychiatric traits between MHQ-responders (N= 118,656) and MHQ-non-responders (N= 243,476) using a z-test. Also, we assessed PRS-distribution differences between both groups using a t-test.

### Within-spouses and within-siblings polygenic risk correlation assessment

In addition to using the GPD approach, we also estimated AM testing PRS correlation within spouses (WSps) and within siblings (WSib) available in the UKB cohort. The first analysis can be informative of the AM in the current generation, while the second one is informative of the AM in the previous generations.

Putative spouses were identified using a method described previously^59^. First, we selected only UKB EUR participants identified as unrelated by kinship coefficients (N=362,132). Then, we selected pairs of individuals who were of opposite sex that reported identical and complete information for the following fields: (a) living with their spouse (UKB field ID: 6141), (b) length of time living in the house (UKB field ID: 699), (c) number of occupants in the household (UKB field ID: 709), (d) number of vehicles (UKB field ID: 728), (e) accommodation type and rental status (UKB field IDs: 670, 680), (f) home coordinates (UKB field IDs: 20074, 20075) and (g) registered in the same UKB recruitment center (UKB field ID: 54) and (h) available genotype data. When more than two individuals shared identical information, then these individuals were removed. To confirm the lack of relatedness in the selected sample, we recalculated the kinship coefficients using Kinship-based INference for GWAS (KING) toolset^60^. Three closely related pairs (identical by descent > 0.1) were removed and only those individuals classified as unrelated by KING were further analyzed. A total of 45,570 putative spouse pairs were identified and included in our analysis. The WSps-PRS correlation was calculated as the coefficient of the regression of PRS of spouse 1 onto PRS of spouse 2 and 20 PCs of spouse 2 to adjust by population stratification:

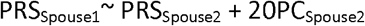

Evidence of possible AM in the current generation was considered when the WSps-PRS correlation was statistically significant after FDR multiple testing correction (FDR q<0.05).

To estimate AM in previous generations, we estimated the WSib-PRS correlation within siblings in the UKB. We selected only UKB EUR participants identified as related by kinship coefficients (n= 64,304). To estimate relatedness in the selected sample, we calculated the kinship coefficients using the KING toolset^60^, and further analyzed only those individuals classified as full siblings by this algorithm. Only two siblings per family were selected for further analysis. A total of 17,911 sibling pairs were included in our analysis.

The WSib-PRS correlation was calculated as the coefficient of the regression of PRS of sibling 1 onto PRS of sibling 2 and 20 PCs of sibling 2 to adjust by population stratification:

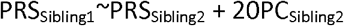

Evidence of possible AM in previous generations was considered significant when the WSib-PRS correlation was statistically different from 0.5 (WSib-PRS rho-0.5Δ) after FDR multiple testing correction (FDR q<0.05).

### Conditional analysis to account for the effect of socioeconomic status on polygenic risk of behavioral and psychiatric traits

To evaluate whether the estimates of AM genetic signatures were affected by SES, we conducted a conditional analysis adjusting the GWAS summary statistics. Specifically, we used the multi-trait-based conditional and joint analysis (mtCOJO)^61^ to adjust UKB-MHQ, PGC, and MVP GWAS statistics by the effect of two SES-related variables: household income (HI, UKB Data-Field 738; i.e., the combined gross income of all members of a household) and the Townsend deprivation index (TDI, UKB Data-Field 189; i.e., a measure of material deprivation based on four variables: unemployment, non-car ownership, non-home ownership, and household overcrowding aggregated for postcodes of residence)^62^. The p-value threshold to select SNPs for clumping in mtCOJO was 0.05. We generated SES-adjusted GWAS summary statistics considering three models: i) HI-adjusted, ii) TDI-adjusted, and iii) adjusted for both HI and TDI. The HI and TDI GWAS statistics were generated by analyzing unrelated EUR UKB-MHQ responders (N=118,656) as described in the Supplementary Methods.

Leveraging the SES-adjusted GWAS summary statistics, the estimates of AM genetic signatures for the behavioral and psychiatric traits were re-estimated and statistical differences between original and SES-adjusted estimates were tested with z-tests. A false discovery rate (FDR q11<110.05) was applied to correct the results for the number of phenotypes evaluated. Furthermore, we evaluated the effect of SES-adjustment on heritability by estimating SNP-h^2^ for the SES-adjusted GWAS summary statistics of the included traits using LDSC^51^.

## Results

### Genetic signatures of assortative mating in MHQ-derived traits

We investigated genetic signatures of AM on MHQ-derived traits (Supplementary Table 2) using three different approaches: GPD^5^, WSib-PRS correlation^63^, and WSps-PRS correlation^59^ (Figure 1). WSib- and WSps-PRS correlation analyses identified multiple AM genetic signatures surviving FDR multiple testing correction, while GPD analysis showed only nominally significant findings. However, although they model different aspects of AM, we observed consistency across the three methods. Interestingly, the MHQ-derived traits showing evidence of AM in at least two analyses were all related to substance use (mostly alcohol consumption) and emotional well-being. The two traits that showed significant estimates in all analyses (WSib- and WSps-PRS correlation FDR q<0.05 and GPD p<0.05) were *Frequency of drinking alcohol* (UKB Field ID: 20414) and General happiness with own health (UKB Field ID: 20459). Consistency between WSib- and WSps-PRS correlation analyses (FDR q<0.05 in both) was also observed for *Amount of alcohol drunk on a typical drinking day* (UKB Field ID: 20403), *Ever taken cannabis* (UKB Field ID: 20453), and *Recent feelings of tiredness or low energy*, UKB Field ID: 20519). The WSps-PR correlation and GPD analyses showed significant results (FDR q<0.05 and p<0.05, respectively) for *Frequency of consuming six or more units of alcohol* (UKB Field ID: 20416) and Felt distant from other people in past month (UKB Field ID: 20496). Below, we described the results obtained in each analysis and the differences observed after adjusting for SES variables.

**Figure 1.**
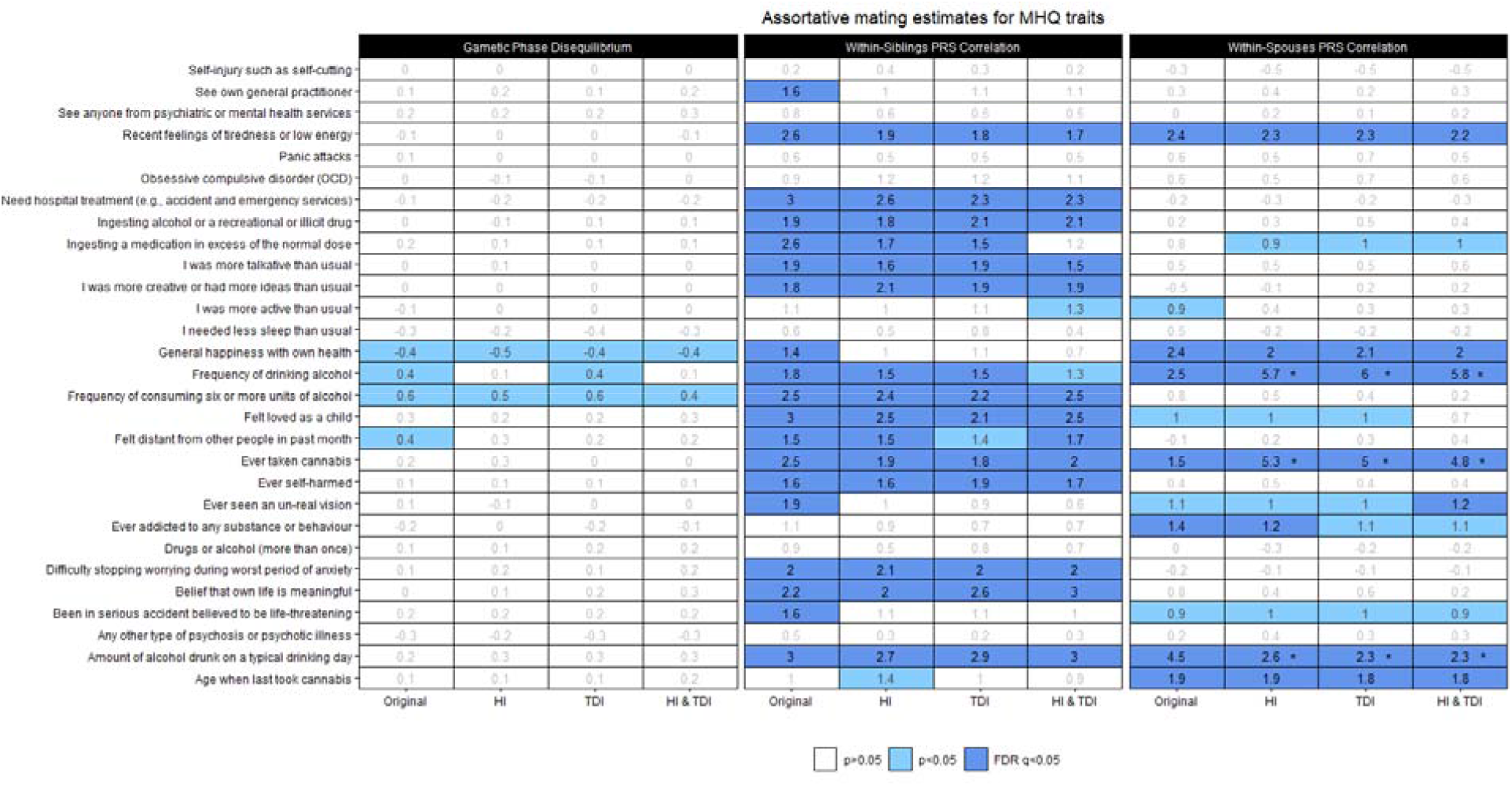
Genetic signatures of assortative mating across Mental Health Questionary (MHQ) traits. Cell shades correspond to the significance strength of each estimate, from white (p > 0.05, non-significant), light blue (p < 0.05, nominally-significant), to dark blue (FDR q < 0.05, FDR-significant). The estimate reported as a percentage is show in the center of each cell. An asterisk in the cell indicates a significant difference of the SES-conditioned estimate with respect to the original estimate (difference-p<0.05). Abbreviations: Household income (HI); Townsend Deprivation Index (TDI).

In the WSib-PRS correlation analysis, we identified FDR-significant results with respect to 19 MHQ-derived traits (FDR q<0.05; Figure 1, Supplemental Table 3). These included substance use (e.g., *Frequency of consuming six or more units of alcohol:* WSib-PRS rho-0.5Δ=0.03, p=7.05×10^−5^, UKB Field ID: 20416; *Ever taken cannabis*: Wsib-PRS rho-0.5Δ=0.03, p=9.14×10^−5^, UKB Field ID:20453), self-harm behaviors (e.g., *Ingesting a medication in excess of the normal dose*: Wsib-PRS rho-0.5Δ=0.03, p=5.41×10^−5^, UKB Field ID:20553, coding:4; *Need hospital treatment following self-harm:* Wsib-PRS rho-0.5Δ=0.03, p=2.14×10^−6^, UKB Field ID:20554, coding:3), negative emotions (e.g., *Recent feelings of tiredness or low energy*: Wsib-PRS rho-0.5Δ=0.03, p=5.55×10^−5^, UKB Field ID:20519), positive emotions (e.g., *Belief that own life is meaningful*: Wsib-PRS rho-0.5Δ=0.02, p=5.31×10^−4^, UKB Field ID:20460), social support (*Felt loved as a child*: Wsib-PRS rho-0.5Δ=0.03, p=2.48×10^−6^, UKB Field ID:20489), mania manifestations (e.g., I *was more talkative than usual*: Wsib-PRS rho-0.5Δ=0.02, p=0.003, UKB Field ID:20548, coding:1), and other psychiatric symptoms (e.g., *Difficulty stopping worrying during worst period of anxiety*: Wsib-PRS rho-0.5Δ=0.02, p=0.002, UKB Field ID:20541). Wsib-PRS rho-0.5Δ estimates remained virtually unchanged after conditioning for SES variables (Figure 1, Supplemental Table 3).

Considering the WSps-PRS correlation, we observed significant results surviving multiple testing correction (FDR q<0.05) with respect to seven MHQ-derived traits (Figure 1, Supplemental Table 4). These included *Ever addicted to any substance or behaviour* (WSps-PRS rho=0.014, p=0.003; UKB Field ID:20401), *Amount of alcohol drunk on a typical drinking day* (WSps-PRS rho=0.045, p=1.29×10^−21^; UKB Field ID:20403), *Frequency of drinking alcohol* (WSps-PRS rho=0.025, p=1.52×10^−7^; UKB Field ID:20414), *Ever taken cannabis* (WSps-PRS rho=0.042, p=0.002; UKB Field ID:20453), *Age when last took cannabis* (WSps-PRS rho=0.019, p=4.11×10^−5^; UKB Field ID: 20455, *General happiness with own health* (WSps-PRS rho=0.024, p=3.04×10^−5^; UKB Field ID:20459), and *Recent feelings of tiredness or low energy* (WSps-PRS rho=0.024, p=2.85×10^−7^; UKB Field ID:20519). Considering the WSps-PRS correlation conditioned with respect to SES variables, four of these traits showed statistically significant differences when compared to the unconditioned estimates (Figure 1; Supplementary Table 4). Statistically significant increases of the WSps-PRS correlation were observed for *Frequency of drinking alcohol* (original estimate 2.5% vs. TDI-adjusted estimate 6%, difference-p=9.44×10^−8^) and *Ever taken cannabis* (original estimate 1.5% vs. HI-adjusted estimate 5.3%, difference-p=7.88×10^−9^). The *Amount of alcohol drunk on a typical drinking day* WSps-PRS correlation from 4.5% in the original estimate changed to 2.3% in the estimate adjusted for both HI and TDI (difference-p=9.13×10^−4^). While these changes were consistent across the different SES conditioning performed (i.e., HI, TDI, and HI+TDI), a significant reduction in *General happiness with own health* WSps-PRS correlation was observed, decreasing from 2.4% in the original estimate to 0.6% when adjusted for HI (difference-p=0.005), but not for the other SES-adjusted traits (difference-p>0.05).

GPD estimates with respect to MHQ-derived traits were not significant after FDR correction (Figure 1, Supplementary Table 5).

With respect to LDSC h^2^ estimates, we observed nominally-significant differences after adjusting by HI and both HI and TDI for *Frequency of drinking alcohol* (original LDSC h^2^=8.3% vs. LDSC h^2^ HI-adjusted=6.7%; difference-p=0.028; original LDSC h^2^=8.3% vs. LDSC h^2^ HI-TDI-adjusted=6.7%; difference-p=0.028; UKB Field ID:20414) and *Ever taking cannabis* (original LDSC h^2^=7.29% vs. LDSC h^2^ TDI-adjusted=5.6%; difference-p=0.021; UKB Field ID:20453) (Supplemental Table 2).

### Genetic signatures of assortative mating in psychiatric traits and disorders

To further expand the breadth of our study, we investigated genetic signatures of AM on psychiatric traits and disorders previously analyzed by large-scale PGC and MVP GWAS (Supplementary Table 1). For three phenotypes, we had information from both MVP and PGC GWAS. The genetic correlation between traits assessed in both PGC and MVP datasets was 1.01 (se=0.17) for anxiety, 0.50 (se=0.15) for PTSD, and 0.92 (se=0.01) for MDD. The limited genetic correlation between PGC-PTSD (excluding UKB) and MVP-PSTD is in line with what previously reported^64^. We investigated GPD estimates in psychiatric traits and disorders in the whole sample combining MHQ responders and non-responders (Supplementary Table 6; Supplementary Figure 1), as well as differences between MHQ responders vs. non-responders with respect to PRS (Supplementary Table 7; Supplementary Figure 2) and GPD for psychiatric disorders. Comparing GPD, WSib-PRS correlation, and WSps-PRS correlation results (Figure 2), we observed FDR-significant AM genetic signatures in at least two different methods for seven phenotypes. In particular, maximum habitual alcohol intake showed FDR-significant results in all approaches and in both MHQ responders and non-responders. Similar consistency was observed for MDD assessed in MVP and Tourette syndrome where FDR-significant evidence was observed in the GPD analysis (both MHQ responders and non-responders) and WSib-PRS correlation with only nominally significant WSps-PRS correlation. Below, we described the results obtained in each analysis and the differences observed after adjusting for SES variables.

**Figure 2.**
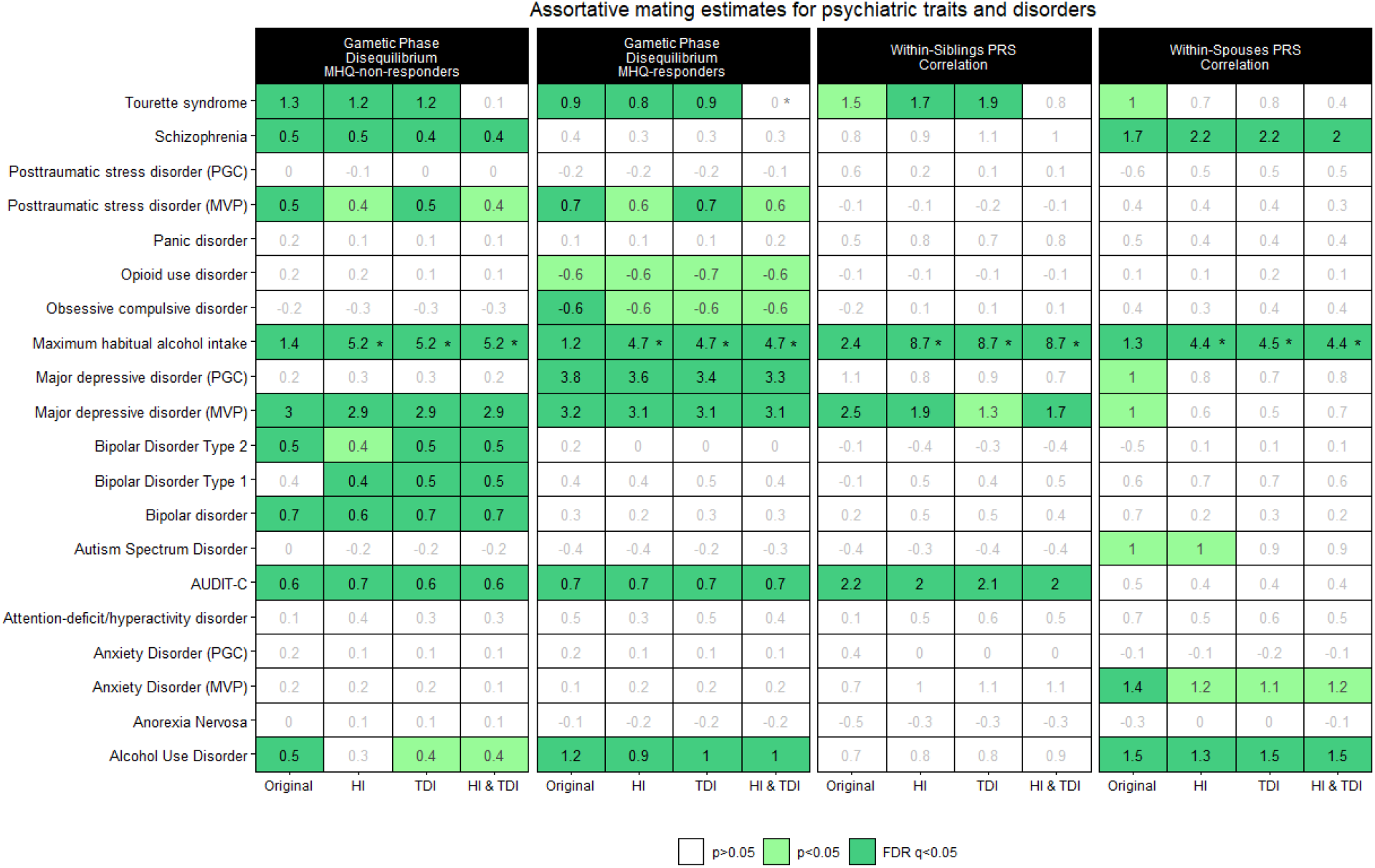
Genetic signatures of assortative mating across psychiatric traits and disorders. Cell shades correspond to the significance strength of each estimate, from white (p > 0.05, non-significant), light green (p < 0.05, nominally significant), to dark green (FDR q < 0.05, FDR-significant). The estimate reported as a percentage is shown in the center of each cell. An asterisk in the cell indicates a significant difference of the SES-conditioned estimate with respect to the original estimate (difference-p<0.05). Abbreviations: Psychiatric Genomics Consortium (PGC); Million Veteran Program (MVP); Mental Health Questionary (MHQ); Household income (HI); Townsend Deprivation Index (TDI).

As mentioned above, the GPD analysis was conducted in MHQ responders and non-responders, separately (Figure 2; Supplementary Tables 8 and 9, respectively). We found significant GPD estimates (FDR q<0.05) for six psychiatric traits in both groups: AUD (GPD_Responders_=1.2%, p=7.09×10^−6^; GPD_Non-Responders_=0.45%, p=0.016), AUDIT-C (GPD_Responders_=0.7%, p=0.008; GPD_Non-Responders_=0.62%, p=7.92×10^−4^), MVP-MDD (GPD_Responders_= 3.23%, p=9.44×10^−33^; GPD_Non-Responders_=2.98%, p=7.04×10^−55^), maximum habitual alcohol intake (GPD_Responders_= 1.22%, p=8.43×10^−6^; GPD_Non-Responders_=1.45%, p=3.99×10^−14^), MVP-PTSD (GPD_Responders_=0.66%, p=0.018; GPD_Non-Responders_=0.52%, p=0.008), and Tourette syndrome (GPD _Responders_=0.94%, p=3.22×10^−4^; GPD_Non-Responders_=1.26%, p=4.56×10^−12^). Also, we found FDR-significant GPD estimates related to BD (GPD_Non-Responders_=0.68%, p=1.69×10^−4^), BD1 (GPD_Non-Responders_=0.39%, p=0.036), BD2 (GPD =0.46%, p=0.016), and schizophrenia (GPD =0.51%, p=9.03×10^−4^) in MHQ-non-responders. While there were significant GPD estimates with respect to PGC-MDD (GPD_Non-Responders_= 3.83%, p=7.5×10^−50^) and OCD (GPD_Responders_=-0.64%, p=0.018) in MHQ-responders. Although there was some variation in the GDP estimates between the two samples investigated, we found significant differences between MHQ-responders and MHQ-non-responders only for PGC-MDD (GPD_Responders_=3.83% vs. GPD_Non-Responders_=0.23%, difference-p=2.27×10^−30^) and AUD (GPD_Non-Responders_=1.2% vs. GPD_Non-Responders_=0.45%, difference-p=0.02). Also, we found a significant difference for PGC-MDD GPD estimates in MHQ-responders (GPD_EUR_=1.44% vs. GPD =3.83%, difference-p=8.36×10^−16^) and MHQ-non-responders (GPD_EUR_=1.44% vs. GPD_Non-Responders_=0.23%, difference-p=1.79×10^−7^) with respect to those in EUR (Supplementary Table 10).

After conditioning on SES variables, we observed statistically significant changes in the GPD estimates only for maximum habitual alcohol intake and Tourette syndrome. These changes were observable in both MHQ responders and non-responders (Figure 2; Supplementary Tables 8 and 9). For TDI adjustment of maximum habitual alcohol intake, GPD increased from 1.22% and 1.45% to 4.74% and 5.21% in MHQ-responders (difference-p=1.61×10^−20^) and MHQ-non-responders (difference-p=3.68×10^−45^), respectively. Conversely, HI-TDI adjustment reduced the GPD estimates for Tourette syndrome from 0.94% and 1.26% in MHQ-responders down to <0.001% and 0.14% (difference-p=0.018) in MHQ-non-responders (difference-p=2.47×10^−5^).

We observed significant WSps-PRS correlation (FDR q<0.05) with respect to four traits (Figure 2, Supplementary Table 11): AUD (WSps-PRS rho=1.47%, p=0.002), MVP-assessed anxiety disorder (WSps-PRS rho=1.4%, p=0.003), maximum habitual alcohol intake (WSps-PRS rho=1.32%, p=0.005), and schizophrenia (WSps-PRS rho =1.74%, p=2.11×10^−4^). Conditioning for SES variables, we observed significant changes in the WSps-PRS correlation only for maximum habitual alcohol intake where the estimate increased from 1.32% to 4.47% after accounting for TDI (difference-p=2.24×10^−6^). A similar SES effect was also present in WSib-PRS correlation analysis where the WSib-PRS rho-0.5Δ estimate changed from 2.4% (p=1.39×10^−4^) to 5.87% (p=1.97×10^−45^) after accounting for TDI (difference-p=8.32×10^−13^). FDR-significant WSib-PRS rho-0.5Δ estimates were observed also for other two traits (Figure 2; Supplementary Table 9): AUDIT-C (WSib-PRS rho-0.5Δ=0.022, p=0.001), and MVP-MDD (WSib-PRS rho-0.5Δ=0.025, p=9.61×10^−5^. The conditioning for SES variables did not change the estimates observed for the other traits (difference-p>0.05; Figure 2, Supplementary Table 12).

Considering the results of the SES conditioning across the three methods applied, we observed a strong and statistically significant increase of the AM genetic signature in the polygenic risk of maximum habitual alcohol intake. Specifically, the adjustment for HI, TDI, and both HI and TDI increased GPD, WSib-PRS rho-0.5Δ, and WSps-PRS rho estimates more than three times (Figure 3).

**Figure 3.**
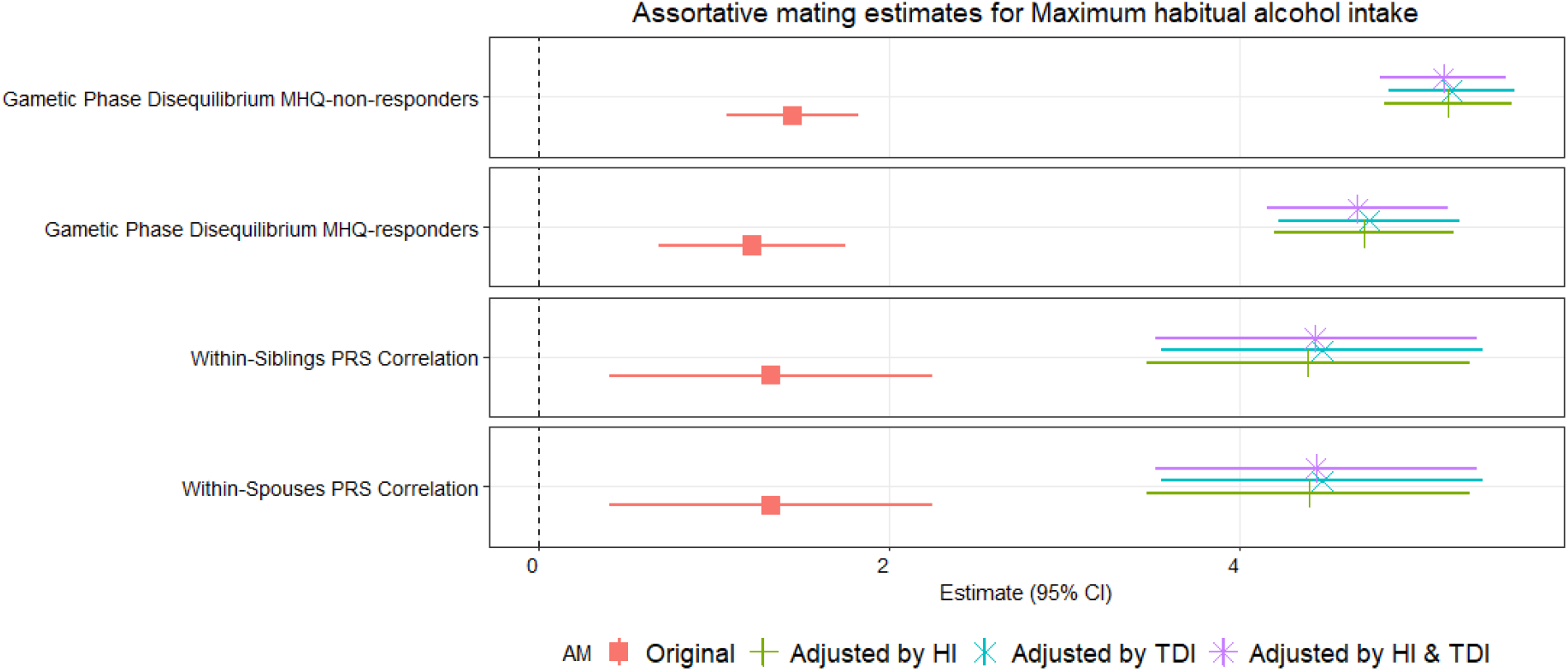
Genetic signatures of assortative mating for Maximum habitual alcohol intake before and after SES conditioning. 95% confidence intervals (CI) are reported for each estimate. Abbreviations: Mental Health Questionary (MHQ); Household income (HI); Townsend Deprivation Index (TDI).

## Discussion

In the present study, we used multiple methods to detect genetic signatures of AM across behavioral and psychiatric traits. We also quantified differences in AM genetic signatures due to i) the effect of SES factors, ii) participation bias by comparing UKB MHQ responders and non-responders, and iii) related to the genetic effect estimates detected by GWAS conducted in samples with different characteristics (i.e., PGC and MVP).

In the UKB MHQ-based analysis, we found consistent evidence of AM genetic signatures across multiple methods for traits related to substance use and emotional well-being. With respect to substance use, three outcomes were related to alcohol consumption (i.e., *Frequency of drinking alcohol, Amount of alcohol drunk on a typical drinking day*, and *Frequency of consuming six or more units of alcohol*) and one to cannabis use (i.e., *Ever taken cannabis*). While several studies have demonstrated phenotypic resemblance between spouses pairs regarding alcohol drinking behaviors^65-67^, limited information is available regarding AM and cannabis use. It has been hypothesized that individuals pick companions compatible with, and supportive of, their substance use, leading to high levels of similarity between romantic partners^68^. Similar mechanisms could be responsible also for the genetic signatures observed with respect to individual feelings (i.e., *General happiness with own health, Recent feelings of tiredness or low energy*, and *Felt distant from other people in past month*). Indeed, evidence of the effect of negative emotions on mating preferences has been previously reported^69^. Interestingly, *General happiness with own health* showed significant evidence of AM across the three methods applied, but with a nominally-significant negative GDP. This suggests that factors other than AM may affect the genetics of this trait in UKB. With respect to possible temporal changes, our results showed mostly evidence of AM both in current and past generations (i.e., significant PRS correlation within spouses and within siblings, respectively). However, considering the two domains identified (i.e., substance use and emotions), *Frequency of consuming six or more units of alcohol, Felt distant from other people in past month, Difficulty stopping worrying during worst period of anxiety*, and *Belief that own life is meaningful* showed genetic signatures for past-generations AM but not for current generation AM. A similar pattern was present also for other MHQ traits, including those related to self-harm (e.g., *Ever self-harmed*) and mania manifestation (e.g., *I was more talkative than usual*). Conversely, the only two traits with evidence of current-generation AM and not to past-generations AM were related to substance use: *Age when last took cannabis* and *Ever addicted to any substance or behaviour*. These different AM patterns may be related to temporal changes in demographic phenomena in the UK populations. For instance, the increased availability of cannabis^70^ may influence mating preferences more in the current generation than in previous ones.

With respect to the SES conditioning in the MHQ analyses, changes in the AM genetic signatures were observed only for traits related to substance use with respect to the WSps-PRS correlation. However, they showed different patterns: increased WSps-PRS correlation for *Frequency of drinking alcohol* and *Ever taken cannabis* and reduced WSps-PRS correlation for *Frequency of consuming six or more units of alcohol* after SES conditioning. This could be due to the known genetic differences between substance use and substance use disorders observed for alcohol, cannabis, and other addictive substances^71-73^. Additionally, misreports and longitudinal changes have been reported to bias genetic associations of alcohol-drinking behaviors^74^. These factors could contribute to the effect of SES conditioning on the WSps-PRS correlation of MHQ-derived alcohol consumption phenotypes together with the complex association of SES observed across the spectrum of alcohol use traits^75^. The analyses based on PGC and MVP GWAS data also showed AM genetic signatures across multiple alcohol-drinking phenotypes: AUD (GPD_responders_; GPD_non-responders_; WSps-PRS), AUDIT-C (GPD_responders_; GPD_non-responders_; WSib-PRS), and maximum habitual alcohol intake (GDP_responders_; GDP_non-responders_; WSib-PRS; WSps-PRS). Interestingly, while significant GDP estimates were observed across these traits, the possible AM temporal scale appears to be different. AUD, an indicator of alcohol addiction, showed AM genetic signatures only in the current generation (i.e., significant estimates for WSps-PRS and not for WSib-PRS). Conversely, AUDIT-C, an indicator of alcohol consumption, showed AM genetic signatures only in the past generations (i.e., significant estimates for WSib-PRS and not for WSps-PRS). In line with the fact that it is less related to addiction than AUD and is more genetically correlated with AUDIT-C than AUD in UKB^76^, maximum habitual alcohol intake showed AM genetic signatures in both current and past generations. Additionally, SES conditioning showed inflated AM genetic signatures of maximum habitual alcohol intake across all methods used (Figure 3). Accordingly, the same population phenomena and/or confounders contributing to the SES-conditioned inflation of AM genetic signature observed in UKB MHQ substance-use traits (i.e., *Frequency of drinking alcohol* and *Ever taken cannabis*) could also be involved in the SES-conditioned inflation of AM genetic signatures observed in MVP maximum habitual alcohol intake. Unfortunately, the limited availability of large-scale GWAS of substance use disorders^77^ did not permit us to fully explore patterns of fully AM genetic differences across different substances.

The analysis of PGC and MVP GWAS data (not including UKB) allowed us to explore GPD differences between UKB MHQ responders and non-responders. While most of the GDP estimates were not statistically different between these UKB subsamples, we observed higher GDP estimates for AUD and PGC-assessed MDD in MHQ responders than in non-responders. As previously described^28,78^, UKB MHQ responders have higher socioeconomic status, are healthier, and report less severe internalizing symptoms. Accordingly, the differences observed may be due to the MHQ participation bias, which may not only affect the generalizability of the prevalence of the MHQ-assessed traits but also the characterization of the polygenic architecture.

Because we had access to PGC and MVP GWAS data related to the same phenotypes (anxiety disorder, MDD, and PTSD), we were able to consider this additional layer of variability. As previously shown^28,79,80^ and confirmed in this study, there is a moderate to high PGC-MVP genetic correlation with respect to these traits. As mentioned, PGC-MDD showed different GPD estimates between MHQ responders and non-responders (3.8% vs. 0.2%). Conversely, MVP-MDD showed FDR-significant and similar GPD estimates in both groups (3.2% vs. 3%). With respect to PTSD, while there was no difference with respect to MHQ participation, FDR-significant GPD was observed when testing MVP-PTSD PRS (responders=0.66%; non-responders=0.52%) but not when considering PGC-PTSD PRS (responders=-0.02%; non-responders=-0.01%). Instead, anxiety disorder showed only FDR-significant WSps-PRS correlation when testing MVP data but not when considering PGC data. These differences are likely due to the characteristics of the PGC and MVP cohorts. PGC GWAS are based on the meta-analysis of many cohorts including participants from different countries that were assessed with different instruments and were enrolled with different recruiting strategies^19,81^. Conversely, MVP GWAS are based on a single cohort that includes only US veterans that were assessed with the same instruments and were enrolled through the same recruiting strategy^20^. Based on the differences observed between PGC-vs. MVP-based analyses, we hypothesize that the analysis of AM genetic signatures based on PRS generated from GWAS meta-analyses may be less affected by the specific characteristics of a cohort (e.g., target population group, assessment, and recruitment strategy). Instead, the analysis of AM genetic signatures based on PRS generated from GWAS conducted in a single cohort may be more affected by the characteristics of that cohort. For example, MVP and UKB cohorts likely present population dynamics that are specific to US and UK demographic histories, respectively^82,83^. Although PRS differences are plausible contributors to GPD differences between both groups, the widespread presence of such differences across the analyzed phenotypes i.e., not being limited to phenotypes where we found significant AM estimates, and the opposite direction of such differences to GPD estimates make them unlikely to significantly influence our results. Accordingly, the AM genetic signatures generated from the analysis of GWAS generated from MVP and UKB-MHQ data could be influenced by cohort-specific characteristics.

Although our study provides new insights into the impact of AM, participation bias, and SES on the polygenic risk of behavioral and psychiatric traits, we acknowledge several limitations. First, due to confidentiality, the identified spouse pairs cannot be confirmed. Thus, it is possible that to an unknown extent the identified pairs do not correspond to actual spouse pairs. Second, the statistical power of the GWAS used to generate the PRS may have contributed to the differences observed across methods and datasets. Third, we cannot discard that observed changes in AM estimates after controlling for SES-related variables may be partially influenced by a potential collider bias^84^. Fourth, the limited availability of large-scale GWAS representative of diverse ancestry groups limited the present study only to data generated from participants of European descent.

In conclusion, we provide evidence of the possible interplay among AM, participation bias, and SES in the polygenic risk of multiple behavioral and psychiatric disorders. Our findings indicate that population phenomena and cohort-specific characteristics could influence our ability to model the polygenicity of traits related to mental health. This highlights the need to model more accurately different aspects that could influence the generalizability of genetic effects detected across cohorts and study designs.

## Data Availability

All results used to make conclusions discussed in this study are provided as Supplementary Material.

## Data availability

All results used to make conclusions discussed in this study are provided as Supplementary Material. All GWAS data are publicly available on their respective websites.

UK Biobank, https://www.ukbiobank.ac.uk/enable-your-research/apply-for-access

Psychiatric Genomics Consortium, https://pgc.unc.edu/for-researchers/download-results/

Million Veteran Program, https://www.ncbi.nlm.nih.gov/projects/gap/cgi-bin/study.cgi?study_id=phs001672.v8.p1

## Acknowledgements

We thank the participants and the investigators involved in the UK Biobank, Million Veteran Program, and the Psychiatric Genomics Consortium for making their data publicly available. This research was conducted using the UK Biobank Resource (application reference no. 58146). The authors acknowledge support from the National Institutes of Health (R21 DC018098, R33 DA047527, RF1 MH132337, and K99 AG078503), One Mind, and the American Foundation for Suicide Prevention (PDF-1-022-21).

