## Supplemental Methods and Figures for "The impact of assortative mating, participation bias, and socioeconomic status on the polygenic risk of behavioral and psychiatric traits"

**Supplementary Methods and Results**

*Heritability estimation*

We used the Linkage Disequilibrium Score Regression method (LDSC) to estimate the single nucleotide polymorphisms based heritability (SNP-h^2^) of each MHQ trait^1^. For this, the genome-wide association study (GWAS) summary statistics of each MHQ trait were formatted as standard LDSC input and analyzed using the 1000 Genomes phase 3 European dataset as LD reference panel^2^. The major histocompatibility complex (MHC) region (chr6: 26–34Mb) was excluded from the analysis. Also, heritability estimates of MHQ traits using the Scalable and Accurate Implementation of GEneralized (SAIGE)^3^ mixed model were obtained from the pan-ancestry genetic analysis of the UKB (Pan-UKB) website (<https://pan.ukbb.broadinstitute.org>)^4^. Those MHQ traits with a SNP-h^2^ p-value < 0.05 and a SAIGE heritability estimate > 0.03 were selected for the estimation of AM.

*Genome-wide association analysis of socioeconomic status-related variables*

We performed GWAS for two measures of socioeconomic status (SES): household income (HI) and the Townsend deprivation index (TDI) separately in 118,656 unrelated UKB participants of European descent who completed the MHQ (MHQ-responders). HI refers to the combined gross income of all members of a household and was assessed via a touchscreen questionnaire completed by UKB participants^5^. This information was collected using a 5-point scale corresponding to the total household income before tax, 1 being less than £18,000, 2 being £18,000–£29,999, 3 being £30,000–£51,999, 4 being £52,000–£100,000 and 5 being greater than £100,000^6^. TDI is a measure of material deprivation based on four variables: unemployment, non-car ownership, non-home ownership, and household overcrowding aggregated for postcodes of residence^7,8^.

GWAS were performed using a logistic regression implemented in PLINK 1.9^9^, including age, sex and the top ten PC to adjust by population stratification. Analyses include only SNPs with the following criteria: minor allele frequencies ≥ 0.01, missingness <0.05, HWE p values >1 × 10−6, and imputation quality score >0.3. LD pruning algorithm implemented in PLINK (r^2^<0.5 considering a window of 50 SNPs). PCs were calculated using the fast PCA approach implemented in PLINK version 2.0^10,11^. LD score regression intercepts for the results of both GWAS were estimated to distinguish polygenic heritability from inflation.

**Supplementary Figures**

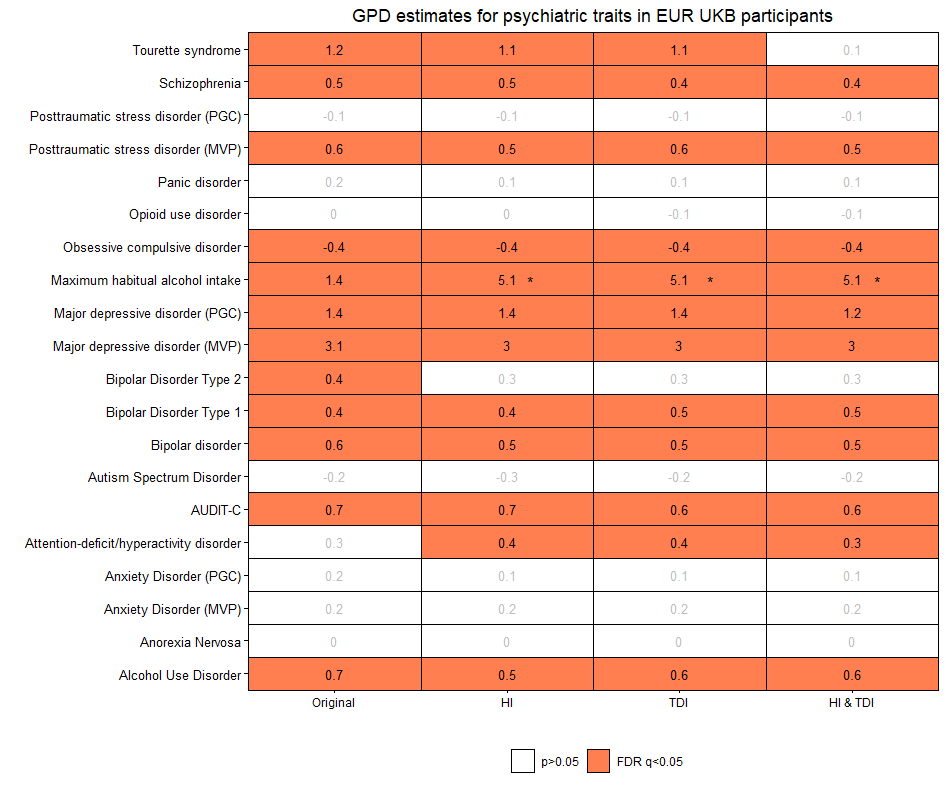

**Supplementary Figure 1.** Genetic signatures of assortative mating across psychiatric traits and disorders in UKB participants of European descent. Cell shades correspond to significance strength of each estimate, from white (p > 0.05, non-significant), to dark coral (FDR q < 0.05, FDR-significant). The estimate reported as a percentage is shown in the center of each cell. An asterisk in the cell indicates a significant difference of the SES-conditioned estimate with respect to the original estimate (difference-p<0.05). Abbreviations: Psychiatric Genomics Consortium (PGC); Million Veteran Program (MVP); Mental Health Questionary (MHQ); Household income (HI); Townsend Deprivation Index (TDI).

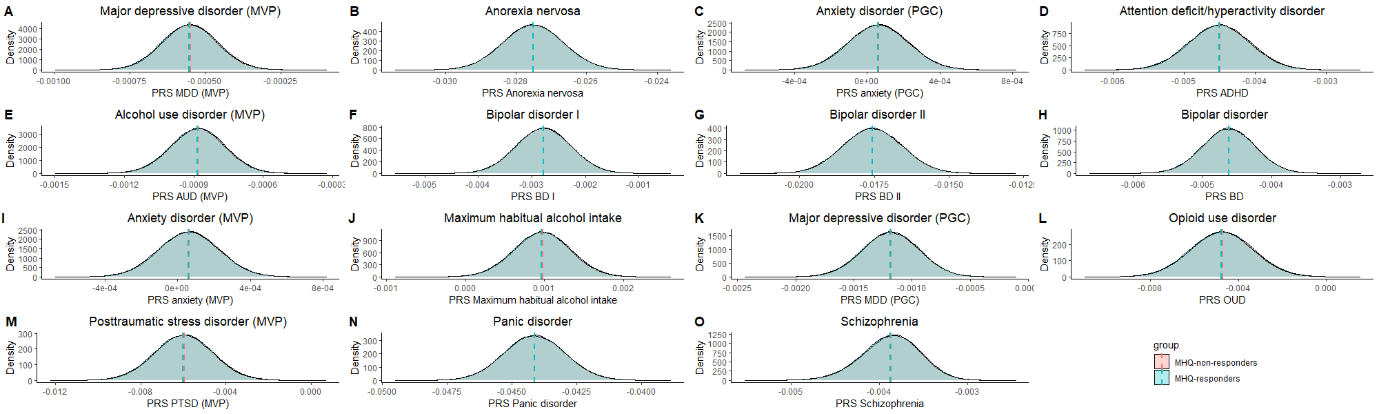

**Supplementary Figure 2.** Polygenic risk score (PRS) distributions in MHQ-responders (blue) and MHQ-non-responders (pink). Mean of PRS distributions is shown as a colored dashed line. Distributions from psychiatric traits with a significant difference (p<0.05) between both groups are shown. Abbreviations: Psychiatric Genomics Consortium (PGC); Million Veteran Program (MVP). Full results are shown in Supplementary Table 7.
